# THE SEARCH FOR AN ASSOCIATION OF HLA ALLELES AND COVID-19 RELATED MORTALITY IN THE RUSSIAN POPULATION

**DOI:** 10.1101/2020.12.22.20248695

**Authors:** Valery Cheranev, Irina Bulusheva, Valery Vechorko, Dmitriy Korostin, Denis Rebrikov

**Affiliations:** Center for Precision Genome Editing and Genetic Technologies for Biomedicine, Pirogov Medical University, 1 Ostrovityanova Street, 117997 Moscow, Russia

## Abstract

HLA genes play a pivotal role in an immune response via the presentation of pathogen peptides in a complex on the surface of cells of a host organism. Here, we studied the association of class I and class II genes with the severity of COVID-19 infection and HLA allele variants.

We performed high-resolution sequencing of class I and class II HLA genes using the sample population of 147 patients who died of COVID-19 and statistically compared our results with the frequencies of the HLA genotypes in a control population of 270 samples.

The obtained data demonstrated that 51:05 and 15:18 alleles from locus B* are statistically significantly associated with COVID-19 severity, while C*14:02 allele correlates with the probability of death from COVID-19 for patients without comorbidities.

## Introduction

In 2020, the human population faced an epidemic that affected almost all countries to a certain degree. It was caused by a new virus from the Coronaviridae family SARS-CoV-2 [1]. The first outbreak of the disease was registered in Wuhan (China) and the disease rapidly spread over the other countries. Currently, the number of registered cases exceeds 53 million, 1.3 million being lethal [2]. In January 2020, the World Health Organization (WHO) declared the outbreak to be a pandemic and termed the disease caused by the virus COVID-19.

One of the essential factors facilitating the rapid spread of COVID-19 is a difference in the severity of a disease and its manifestations widely ranging in individual patients [3, 4, 5]. The accumulated data on SARS-CoV-2 suggest some correlation between the disease outcome and sex, age, and concurrent diseases [6]. Disease severity can be also associated with individual genetic characteristics of a patient [7, 8]. One of the genetic predictors for severity and an outcome of the disease could be class I and II HLA genes encoding the proteins of the major histocompatibility complex (MHC). The main function of these molecules is presenting an antigen on the surface of the plasma membrane for their recognition by immune cells. Therefore, the alleles encoding the aminoacid sequence in MHC directly affect the susceptibility to certain diseases. Based on the data on the alleles of class I and II HLA genes and the protein composition of SARS-CoV-2, we analyzed the level of the affinity of MHC binding to all possible viral epitopes [9]. The lowest predicted level of the interaction with viral antigens belonged to the protein encoded by B*46:01 allele while the highest level belonged to the B*15:03 allele. Thus, we proceeded with a retrospective analysis aimed to reveal any association between the identified alleles and disease severity [10-15].

Previously, the correlation between HLA alleles and the disease in the test and control groups was performed using statistical analysis by the 2-level procedure of identification of significant alleles [16]. On the first stage, the distribution of all alleles in patients was compared to the distribution of the reference population in order to choose several candidate alleles. After that, candidate alleles were compared between two groups of patients using the Bonferroni method.

The correlation between HLA and the disease was studied at the background of concurrent diseases [17, 18, 19].

In our work, we compared the frequencies of different alleles of class I (A, B, C) and class II (DRB1, DQB1) HLA genes between the groups of patients and healthy donors. The main goal of the work is to identify and validate the alleles significantly related to various COVID-19 outcomes.

## Background

### Materials and methods

#### Ethics statement

This study conformed to the principles of the Declaration of Helsinki. The appropriate institutional review board approval for this study was obtained from the Ethics Committee at Pirogov Medical University. All patients provided written informed consent for the collection of samples, subsequent analysis, and publication thereof.

#### Biomaterial collection

The biomaterial consisting of whole venous blood was collected in EDTA-coated tubes used as a preservative.

#### gDNA isolation

gDNA was isolated from 100 ul of whole venous blood with Proba-Mch-Maks reagent kit using automated dosing station дТстрИм (DT stream) (DNA Technology, Russia). Quality check of the isolated DNA was performed using gel electrophoresis in agarose gel, the concentration was measured using Qubit 2 fluorimeter with Qubit dsDNA BR Assay kit (ThermoFisher Scientific, USA).

#### HLA library preparation

The preparation of amplicon libraries for HLA high-resolution genotyping was performed using HLA Expert kit (DNA Technology, Russia), following the manufacturer’s protocol.

#### Sequencing

Sequencing was performed with Illumina MiSeq using MiSeq Reagent Kit v3 (600-cycle), according to the manufacturer’s instructions.

#### HLA high-resolution genotyping

The analysis of fastq files was performed with HLA-Expert software v.2.0 (DNA Technology, Russia) according to the manufacturer’s protocol.

#### Statistical Analysis

Statistical analysis was performed using Arlequin v.3.5.2.2 [16] and Pearson’s goodness-of-fit test. We created several scripts allowing for estimating the diversity of each gene and differences in frequencies of the individual alleles in groups. We also created a script that cleared an input table containing patients’ data from errors and transformed the names of HLA alleles, according to a unified syntax (https://github.com/genomecenter/HLA_article). We created a script that generated an input file containing patients’ data for Arlequin.

## Results

As a study population, we used a collection of venous blood samples and clinical data from 147 patients (group III) that had died in Moscow clinics from coronavirus disease COVID-19 during the pandemic in 2020. The positive test for COVID-19 was confirmed using qPCR kits in Moscow clinical diagnostic laboratories that collected the biomaterial. Clinical features of the group III is presented in Table 1. As the control sample population, we used 270 venous blood samples collected from the members of the National Registry of Bone Marrow Donors by the Pirogov Medical University at the beginning of 2020. All members of the control sample population were surveyed and stratified into two groups: those who hadn’t suffered from COVID-19 during our study (245 samples, group 1) and those who had recovered from COVID-19.

**Table 1.** Clinical data in the cohort of deceased patients with COVID-19.

|  | Age 65 years and below | Above 65 years old | No full clinical presentation |
| --- | --- | --- | --- |
| The number of patients | 30 | 67 | 50 |
| Age, median, Q25-Q75 | 58 (52.5-62.75) | 81 (74.5-86) | - |
| **Sex** |  |  |  |
| Female | 15 | 42 | - |
| Male | 15 | 25 | - |
| **Comorbidities** | 24 | 59 | - |
| Acute respiratory infection | 1 | 0 | - |
| Pneumonia in the absence of respiratory failure | 3 | 6 | - |
| ARDS | 13 | 20 | - |
| Sepsis, septic shock | 5 | 3 | - |
| Thromboses, embolism | 10 | 17 | - |
| Skin rashes | 0 | 0 | - |
| DIC | 0 | 0 | - |
| Multiple organ failure | 12 | 13 | - |
| Chronic inflammatory infectious diseases | 2 | 15 | - |
| Autoimmune diseases including type 1 diabetes mellitus | 0 | 1 | - |
| Metabolic diseases including type 2 diabetes mellitus | 10 | 11 | - |
| Cardiovascular diseases | 17 | 51 | - |

The data from HLA high-resolution genotyping for each patient included the information on both alleles of A*, B*, C*, DRB1*, DQB1* genes in the HLA histocompatibility complex (Group I-III HLA genotypes are presented in Supplemental Table 1). We performed this analysis for every gene and for each allele of a gene that had significant differences. The distribution of allele frequencies over 5 loci from three groups is presented in Figure 1.

**Figure 1.**
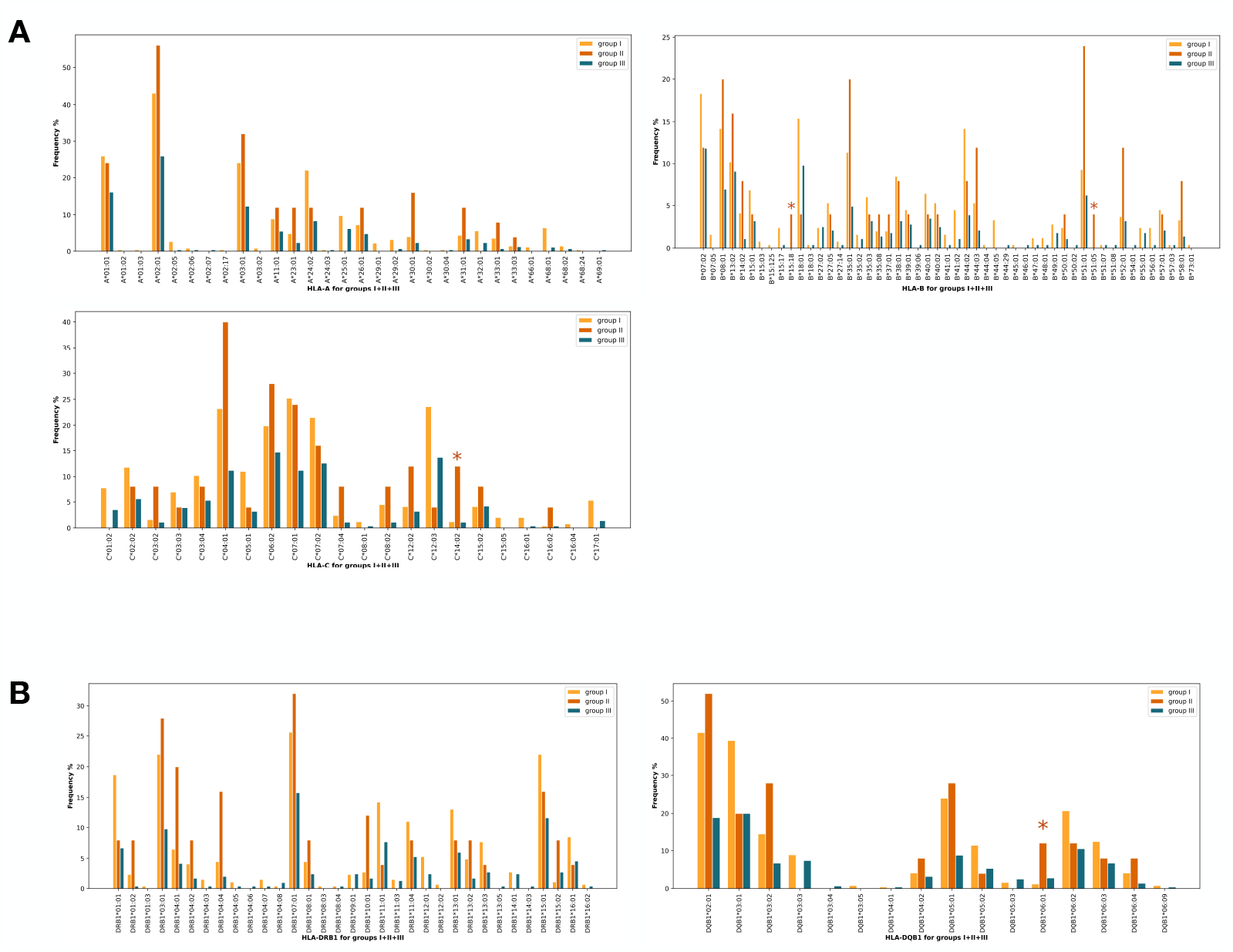
**The histogram of the frequencies of alleles of class I (A) and class II (B) HLA loci. The horizontal axis, the value of the 1st level allele. The vertical axis, its frequency in a sample population. Group I is denoted by yellow bars, group II is denoted by orange bars, group III is denoted by green bars. Starts denote the alleles that showed statistically significant associations**.

For three groups, we used a method of distances upon pairwise comparison [21] using the fixation index FST which is a special case of the Wright F-statistics based on the correlation of randomly chosen alleles within a population with the whole population which is equivalent to the ratio of genetic diversity due to the differences in allele frequency in populations [22]. Results are presented in Table 2. Zero and negative values of FST mean that there was no genetic stratification between populations Group I and III are similar, group II is equidistant from both of them.

**Table 2.** The results obtained by the distance method upon pairwise comparisons of groups I-III.

| Group | I | II | III |
| --- | --- | --- | --- |
| I | 0 |  |  |
| II | 0.00335 | 0 |  |
| III | -0.00045 | 0.00324 | 0 |

After that, we performed the population Assignment Test (Figure 2). For each sample population, we created a probability graph of the genotype within the sample from the population compared to the other populations. The results obtained are similar to those obtained from a pairwise comparison.

**Figure 2.**
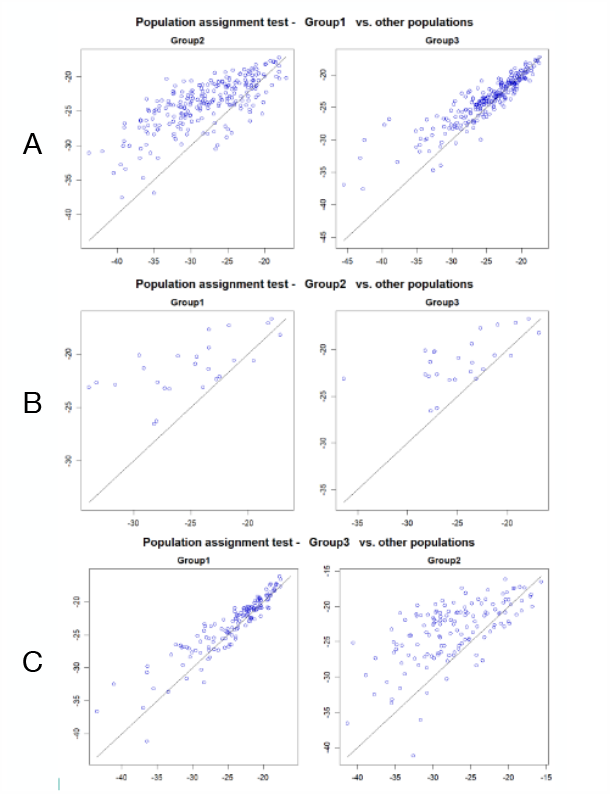
**Results from the population Assignment Test. For each group (I – A, II – B, III – C), we created the genotype probability plot within the sample from the population compared to all other populations**.

Alternatively, we used Pearson’s goodness-of-fit test that enabled a separate analysis of each gene.

We analysed the 2-level alleles for each gene. The null hypothesis stated that HLA did not affect the divergence in the groups. We chose a p-value of 0.05 as a threshold value. We compiled a contingency matrix for each gene using one of four methods (Table 3). Although considering both alleles from each patient as independent ones (V2) is incorrect from the point of view of statistical analysis, we decided to perform this analysis in order to compare the results.

**Table 3.**
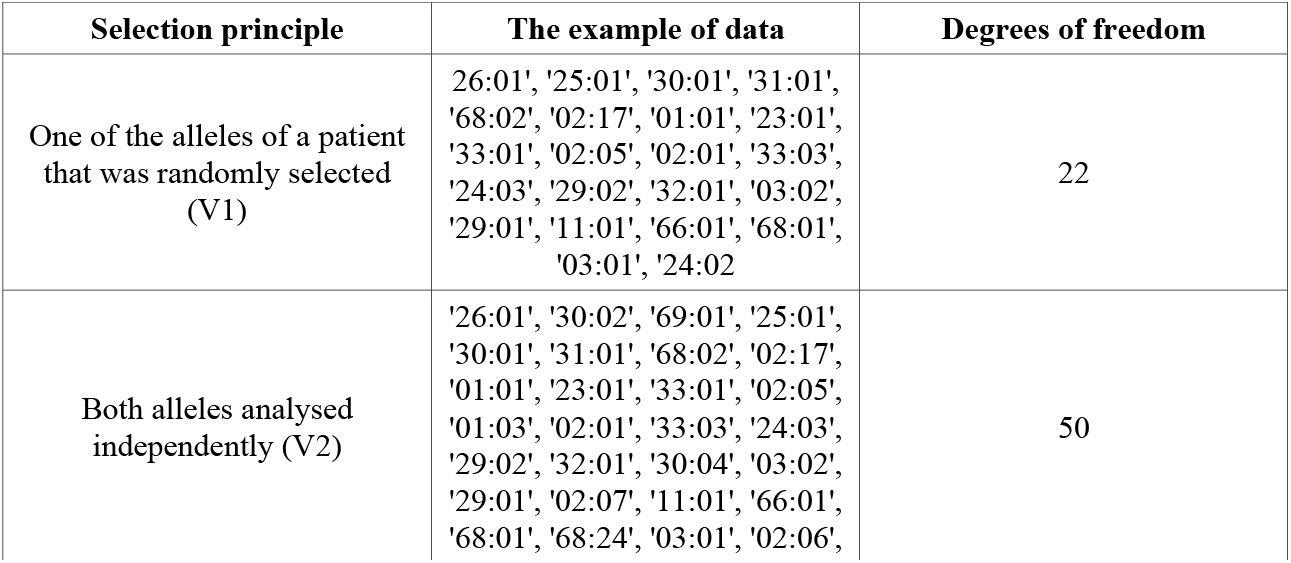
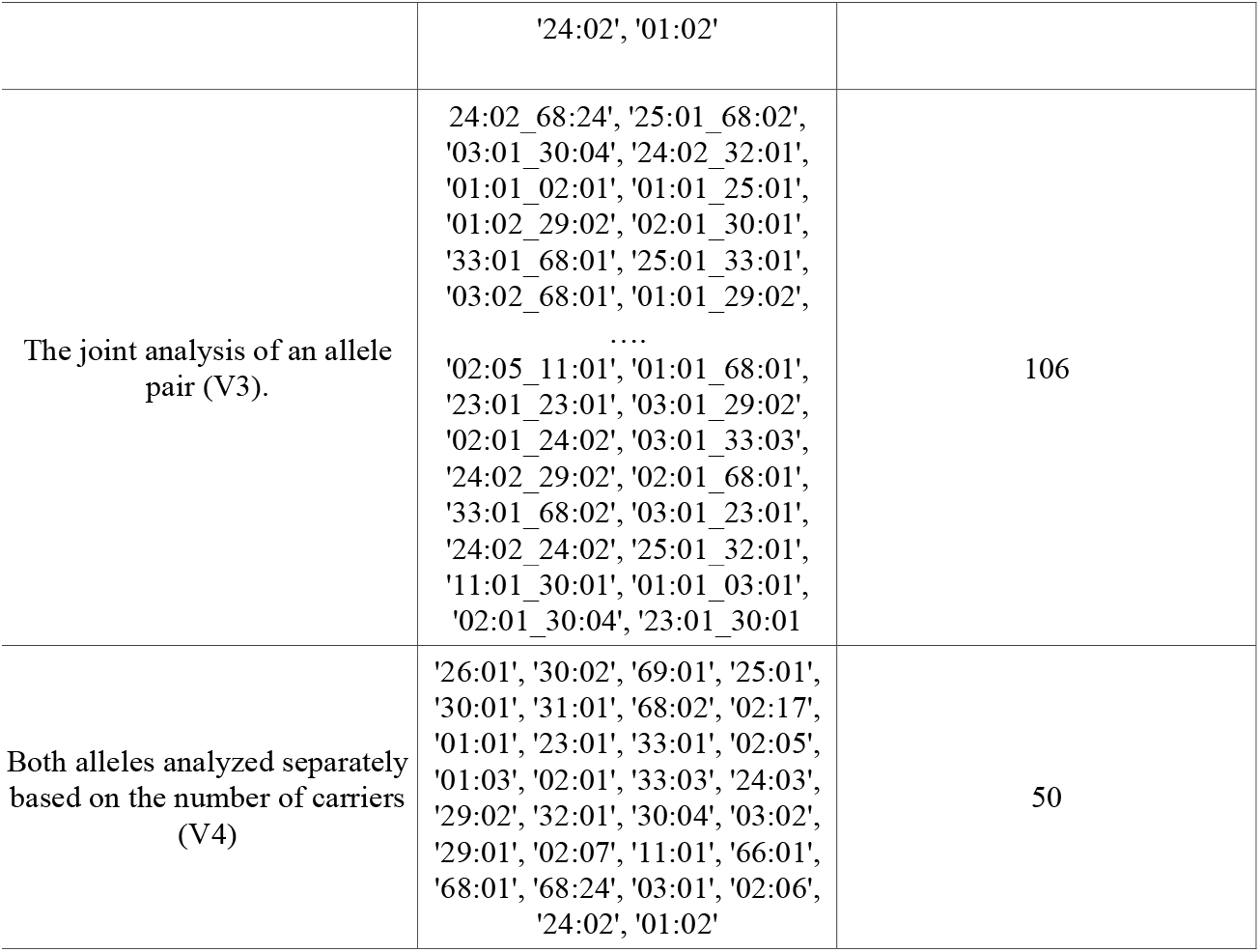
Three ways to collect patient’s data to compile a contingency matrix for each gene.

Analysis of the combined groups: I+II vs III (i.e. healthy or recovered donors vs patients who died from COVID-19), II+III vs I (i.e. all who suffered from COVID-19 vs healthy donors) didn’t reveal any significant differences. Separate analysis of three groups revealed statistically significant effects of HLA-C* allele combinations (Table 4). As a multiple comparison correction, we used the Bonferroni method [23] with the significance level of 0.05 Locus C* significantly affects the group stratification. To determine significant alleles for each gene based on the way of compiling a contingency matrix, we created a special script. The results obtained for V3 are presented in Table 5. For alleles we considered only once, we chose the V4 method, as the results from the V1 method should be considered in the iterative dynamics due to the random choice of an allele. The results obtained from V4 are presented in Table 6.

**Table 4.**
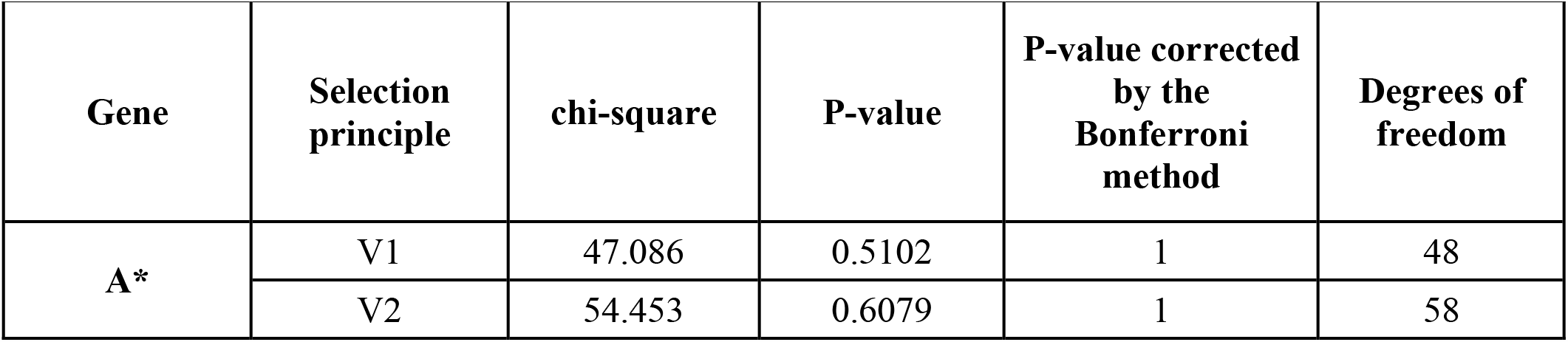
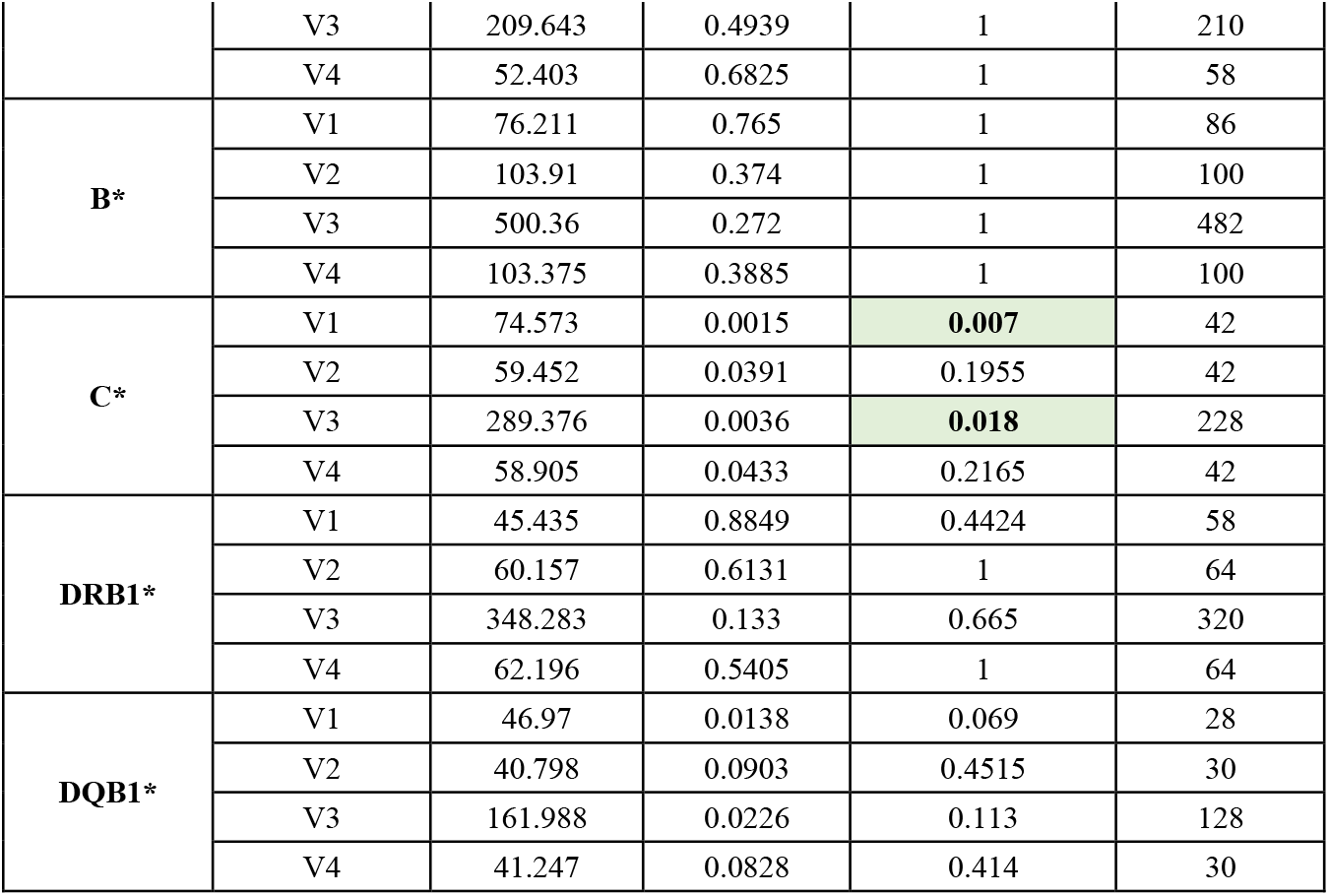
Comparison of groups I, II, and III of samples based on Pearson’s goodness-of-fit test.

**Table 5.** **The list of statistically significant combinations of HLA gene alleles that define the differences between groups I, II, and III following the V3 method**.

| **Locus** | **The significant allele combination** | **P-value corrected by the Bonferroni method** |
| --- | --- | --- |
| **A\*** | 30:01_33:03 | 0.0409 |
|  | 11:01_23:01 | 0.0409 |
| **B\*** | absent | 1 |
| **C\*** | 04:01_14:02 | 0.0444 |
|  | 07:04_14:02 | 0.0444 |
|  | 07:02_16:02 | 0.0444 |
|  | 04:01_08:02 | 0.0444 |
|  | 07:02_07:04 | 0.0444 |
|  | 03:03_14:02 | 0.0444 |
| **DRB1\*** | 04:01_11:04 | 0.0075 |
|  | 04:04_07:01 | 0.0075 |
| **DQB1\*** | 03:02_03:02 | 0.0251 |
|  | 02:01_06:01 | 0.0251 |

**Table 6.**
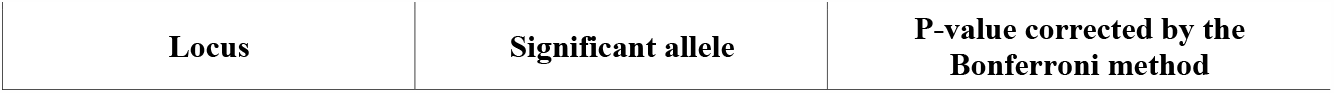
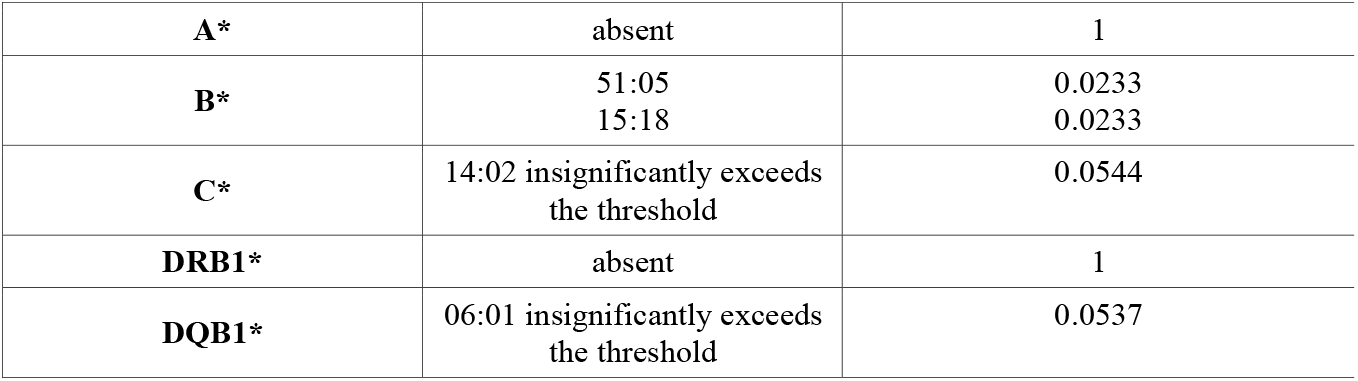
**The list of statistically significant combinations of HLA gene alleles that define the differences between groups I, II, and III following the V4 method**.

To determine the significant alleles, we analysed the other group combinations, according to the V4 method. In particular, the group of deceased patients was divided into subgroups, according to their age (age not exceeding 65 or exceeding 65 at the time of death) and comorbidities. The group designations and sizes are presented in Table 7. The results obtained from the other combinations are presented in

**Table 7.** The designations and number of the groups subjected to analysis.

| **Group name** | **Designation** | **Number** |
| --- | --- | --- |
| Bone marrow donors who didn't suffer from COVID-19 | I | 245 |
| Bone marrow donors who had COVID-19 | II | 25 |
| Patients who died from COVID-19 | III | 147 |
| Patients who died from COVID-19 under age of 65. | IIIA | 30 |
| Patients who died from COVID-19 above age of 65 | IIIB | 67 |
| Patients who died from COVID-19 with comorbidities | IIIC | 83 |
| Patients who died from COVID-19 without comorbidities | IIID | 14 |

Table 8. We should note that we had to exclude some patients from the sample population (Table 1) due to the lack of full clinical data. The distribution of allele frequencies of the locus C* in the subgroups IIIA-IIID is presented in Figure 3.

**Table 8.** Comparison of the group combinations analyzed by V4 method.

| **Group combination** | **Locus** | **Significant allele** | **P-value corrected by the Bonferroni method** |
| --- | --- | --- | --- |
| **(I+II)+III** | - | - |  |
| **I+(II+III)** | - | - |  |
| **I+II+IIIA+IIIB** | C* | 14:02 | 0.03439 |
| **II+IIIA+IIIB** | - | - |  |
| **IIIA+IIIB** | - | - |  |
| **(I+II)+IIIA+IIIB** | DQB1* | 03:04 insignificantly exceeds the threshold | 0.0593 |
| **I+II** | - | - |  |
| **II+III** | - | - |  |
| **IIIC+IIID** | - | - |  |
| **IIIA+IIIB+IIIC+IIID** | C* | 14:02 | 0.0047 |
| **II+IIIC+IIID** | C* | 14:02 insignificantly exceeds the threshold | 0.0637 |
| **I+II+IIIC+IIID** | - | - |  |

**Figure 3.**
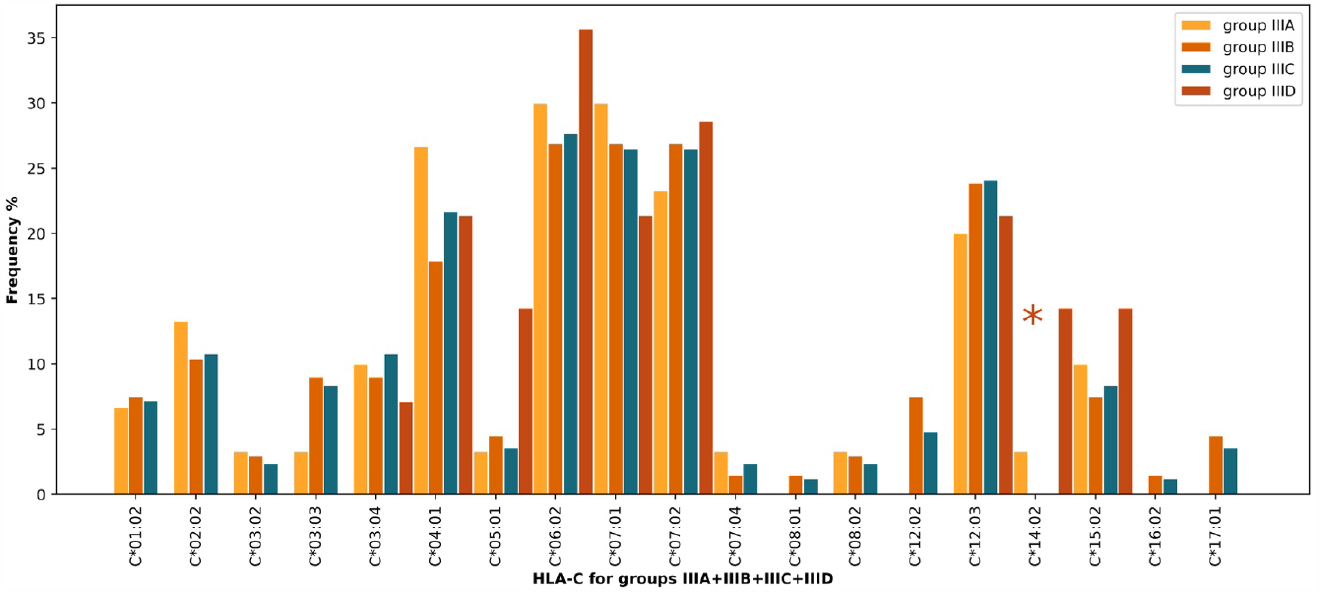
**The distribution of allele frequencies of locus C* in the patients who died from COVID-19. The subgroups are indicated in the diagram**.

## Discussion

Thus, locus C* statistically significantly affects the groups of patients who didn’t suffer from COVID-19, recovered and died (Table 4); estimating the significance of allele combinations reveal statistically significant associations with COVID-19 severity (Table 5). Locus B 51:05 and 15:18 alleles show statistically significant association with the COVID-19 severity (Table 6), while the p-value of C*14:02 and DQB1*06:01 alleles are close to the threshold value. Upon stratification of the patients who died from COVID-19 according to their age, C*14:02 allele was significant (Table 8).

Importantly, the manipulations with group stratification based on various parameters allow determining a criterion that would make the stratification significant. This is related to a limited size of the experimental sample population. Therefore, we are planning to confirm the results obtained in this work using a larger sample that is being collected at the moment. We hope to obtain conclusive evidence regarding the C*14:02 and DQB1*06:01 alleles.

We performed a comparative analysis of the previously published results on the search for the association between HLA and COVID-19 severity (Table 9). Statistically significant and close to statistically significant B*51, C*14:02, and DQB1*06:01 alleles we singled out in our work turned out to be identified in other works as well. Some divergences in the results may be accounted for distinct frequencies of HLA alleles within the particular ethnic groups.

**Table 9.**
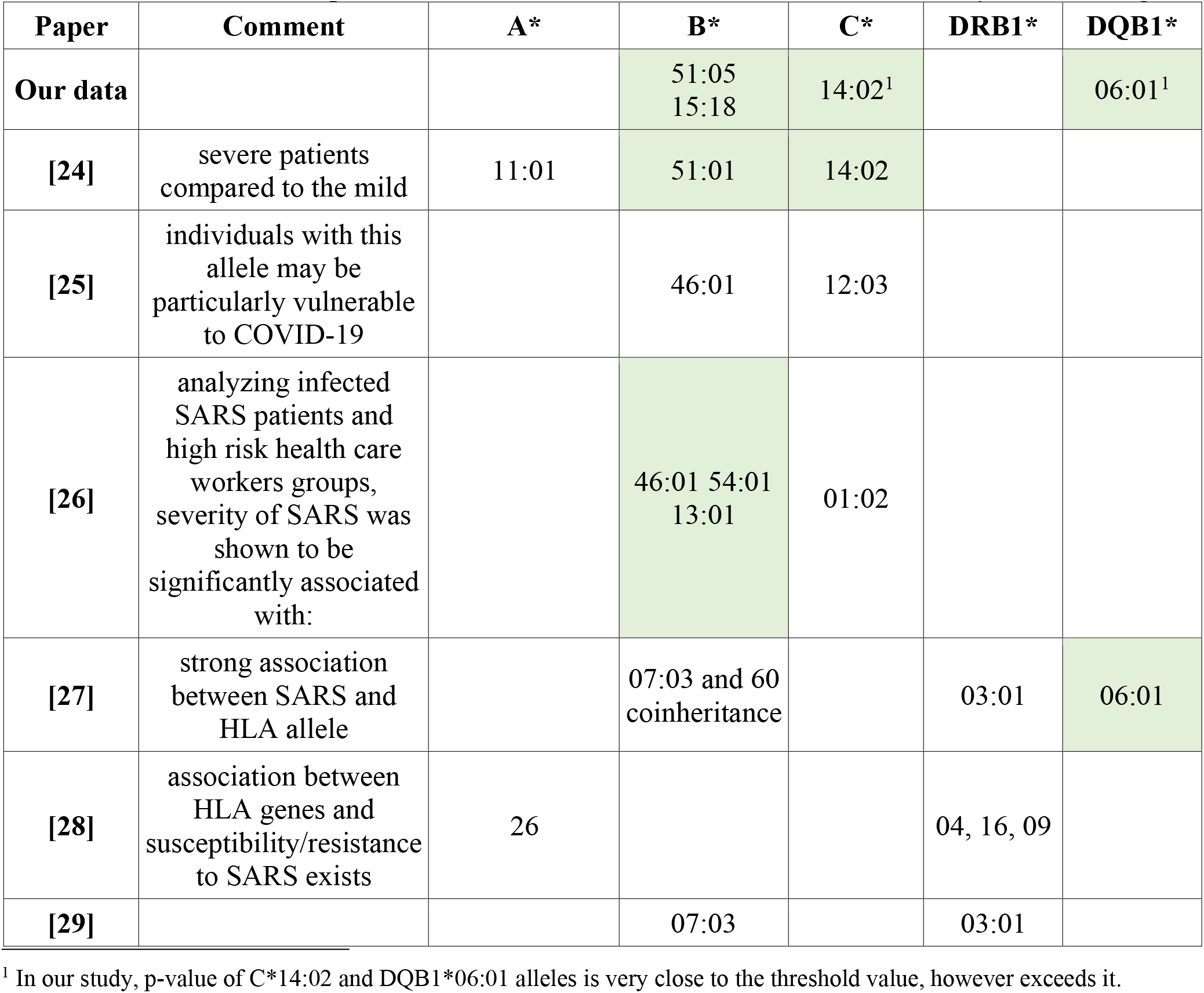
Comparison of the results on alleles for COVID-19 obtained by other investigators.

## Conclusions

Locus B*51:05 and 15:18 alleles show statistically significant association with COVID-19 severity upon comparing the sample populations of patients in Russia who died from coronavirus infection with the group of bone marrow donors. They are more frequent in patients who recovered from COVID-19. The C*14:02 allele revealed a statistically significant correlation with the increased probability of death from COVID-19 of patients without comorbidities.

## Data Availability

All received data are available upon request

